# Measuring Baseline Health with Individual Health-Adjusted Life Expectancy (iHALE)

**DOI:** 10.1101/19003814

**Authors:** Kjell Arne Johansson, Jan-Magnus Økland, Eirin Krüger Skaftun, Gene Bukhman, Ole Frithjof Norheim, Matthew M. Coates, Øystein Ariansen Haaland

## Abstract

**Objectives:** At any point of time, a person’s baseline health is the number of healthy life years they are expected to experience during the course of their lifetime. In this article we propose an equity-relevant health metric, illness-specific individual Health Adjusted Life Expectancy (iHALE), that facilitates comparison of baseline health for individuals at the onset of different medical conditions, and allows for the assessment of which patient groups are worse off. A method for calculating iHALE is presented, and we use this method to rank four conditions in six countries according to several criteria of “worse off” as a proof of concept.

**Methods:** iHALE measures baseline health at an individual level for specific conditions, and consists of two components: past health (before disease onset) and future expected health (after disease onset). Four conditions (acute myeloid leukemia (AML), acute lymphoid leukemia (ALL), schizophrenia, and epilepsy) are analysed in six countries (Ethiopia, Haiti, China, Mexico, United States and Japan). Data for all countries and for all diseases in 2017 were obtained from the Global Burden of Disease Study database. In order to assess who are the worse off, we focus on four measures: the proportion of affected individuals who are expected to attain less than 20 healthy life years (T20), the 25^th^ and 75^th^ percentiles of healthy life years for affected individuals (Q1 and Q3, respectively), and the average iHALE across all affected individuals.

**Results:** Even in settings where average iHALE is similar for two conditions, other measures may vary. One example is AML (average iHALE=58.7, T20=2.1, Q3-Q1=15.3) and ALL (57.7, T20=4.7, Q3-Q1=21.8) in the US. Many illnesses, such as epilepsy, are associated with higher baseline health in high-income settings (average iHALE in Japan=64.3) than in low-income settings (average iHALE in Ethiopia=36.8).

**Conclusion:** iHALE allows for the estimation of the distribution of baseline health of all individuals in a population. Hence, baseline health can be incorporated as an equity consideration in setting priorities for health interventions.

## INTRODUCTION

All health systems have budget constraints and limited resources. Methods for health economic evaluations, like cost-effectiveness analysis (CEA), are essential in health policy and are extensively used to rank health services by their expected efficiency [1]. However, few people endorse strict health maximisation [2], and fairness criteria may be included in such rankings [3, 4]. For example, one may give higher priority to interventions that target those with the most severe illnesses [5-7], especially in relation to decisions about the pricing and reimbursement of new medicines and devices [8, 9]. Policy makers in countries like Norway [10] and the Netherlands [11] have already started using severity measurement methods.

In this paper, the terms “illness”, “disease” and “condition” are used interchangeably, and include all adverse medical conditions, such as injuries, syndromes, birth defects, and infections. The term “severity of illness” involves both substantial value disagreements and a wide range of interpretations [12, 13]. To sidestep misunderstandings, we use the concept of “baseline health at disease onset” rather than severity of illness. Three perspectives on how to measure baseline health dominate in the literature. One view considers current health [14], one considers health over future years [15], and one considers health over the lifetime [16-18]. In this paper we conform to the latter, and focus on how a particular illness is expected to affect the total lifetime health achieved by an individual before they die [18]. This includes both the past (before disease onset) and future (after disease onset). Clinical definitions of severity of illness often include urgency, but our definition of baseline health does not. Urgency pertains to the timing of treatment and how this influences the prognosis of a condition. Conditions with a low baseline health at onset, like multiple sclerosis in young patients, do not necessarily require urgent interventions.

The Global Burden of Disease (GBD) study provides critical summary measures of population health that are relevant when evaluating and comparing health systems [3]. These measures include disability-adjusted life years (DALY) and healthy life expectancy (HALE). DALYs are calculated for a set of diseases by summing the years of life lost (YLL) compared to a reference life expectancy and years lived with disability (YLD) in one particular year due to each disease [19]. For a particular condition and a particular year, YLL is the sum of all the years lost for the individuals who died from the condition during that year. The reference is the age-adjusted life expectancy (LE) from a life table derived from the mortality rates in the locations with the lowest age-specific mortality in the GBD study [20]. YLD, on the other hand, is the sum of the health loss due to the condition during the year across people living with the condition [21]. DALYs aggregated from YLLs and YLDs are a measure of overall population burden. A major limitation of these measures is that they do not capture how the condition affects the distribution of baseline health at onset across individuals in the population.

We propose a framework where this distribution is an integral part. A key component in this framework is the new metric individual Health Adjusted Life Expectancy (iHALE). In this paper, we present a method for calculating iHALE, and how to use iHALE to rank conditions according to baseline health at onset. We consider four conditions and six countries to illustrate how and why our framework is relevant for priority setting in health care and the measurement of population health.

## METHODS

### Definition of iHALE

iHALE measures lifetime health at an individual level for individuals with specific conditions, and consists of two components: past health and future expected health. We obviously do not know the actual time of death for people dying in the future, but we do have some knowledge about the expected distribution across individuals. Consider, for example, two people aged 30 (Ann) and 50 (Bob) who each get a disease. The prognosis for Ann is that she will certainly die within 20 years, but we do not know exactly when. The risk of dying is 99% before her 50^th^ birthday, but there is also a 1% chance that she will die in her 51^st^ year. Bob, however, will certainly die before he is 51. For simplicity, we disregard health/disability adjustment for time with illness, and focus only on their age at death. In a lifetime health perspective, there is a 99% probability that Ann will die at a younger age than Bob will.

Hence, Ann’s baseline health is less than Bob’s in terms of total length of life (past life plus expected future life), even though Bob’s expected future life is shorter. This is true even if there is a 1% chance that Ann too will die in her 51^st^ year. Health adjustment complicates matters, as we will discuss below, but the principles are the same. Of course, iHALE needs to go beyond hypothetical two-person cases to become a relevant health metric for priority setting in countries with millions of individuals and multiple diseases. iHALE enables comparison of both average baseline health and distribution of baseline health between individuals with different diseases. Methods for calculating the iHALE distribution within disease conditions are presented in the next sections.

### Data

For illustrative purposes, we consider four conditions (acute myeloid leukemia (AML), acute lymphoid leukemia (ALL), schizophrenia, and epilepsy) in six countries (Ethiopia, Haiti, China, Mexico, United States and Japan). The diseases have distinct properties that highlight certain characteristics of iHALE. The two leukemias are fatal, but the incidence of ALL peaks at both young and older age groups, whereas AML incidence peaks at old age groups only. Schizophrenia has large impact on disability over many years, and there are variations in both mortality and morbidity of epilepsy across countries. We consider the leukemias in a US setting, and then we compare schizophrenia and epilepsy across the six countries, representing low-, middle- and high-income settings with different age distributions, levels of health systems development, and access to healthcare among their populations. Data for all countries and for all diseases in 2017 are obtained from the freely available GBD cause of death database [20]. Table 1 describes variables available in the GBD database, and how they are used to derive other important variables.

**Table 1:** Description of data and variables used to calculate individual Health Adjusted Life Expectancy (iHALE), the GBD 2017 study [3] is source for all calculations.

| Variable | Description |
| --- | --- |
| PIM | *Period of increased mortality.*<br>From expert opinions.<br>Number of years with increased mortality after disease onset. The rate at which mortality declines in the PIM is specific for each condition.<br>For simplicity, PIM=100 for chronic diseases. We use PIM=5 for the leukemias. |
| PID | *Period of increased disability.*<br>From expert opinions.<br>Number of years with increased disability after disease onset. The rate at which disability declines in the PID is specific for each condition.<br>For simplicity, PID=100 for chronic diseases. We use PID=5 for the leukemias. |
| pop | *Population size, per 5-year age interval.*<br>From GBD 2017.<br>Transformed to 1-year age intervals by distributing individuals evenly across the five years. |
| $P_D$ | *Prevalence (per population) of disease per 5-year age interval.*<br>From GBD 2017.<br>Assumed to be the same in all 1-year intervals. |
| $I_D$ | *Incidence (per population) of disease per 5-year age interval.*<br>From GBD 2017.<br>Assumed to be the same in all 1-year intervals. |
| $M_D$ | *Disease specific probability of death per 5-year age interval, for total population.* |
|  | From GBD 2017.<br>Assumed to be the same in all 1-year intervals. |
| q | *All cause probability of death per 5-year age interval, for total population (background mortality).* From GBD 2017.<br>$q = M_{\text{All causes}}$<br>Assumed to be the same in all 1-year intervals. |
| YLD<sub>D</sub> | *Years Lived with Disability of disease per 5-year age interval.*<br>From GBD 2017.<br>Assumed to be the same in all 1-year intervals. |
| **Derived** |  |
| em<sub>D</sub> | Excess mortality due to disease (case fatality rate). These are not given directly in GBD, but can be calculated using<br>$em_D(\text{age}) = \frac{M_D(\text{age})}{P_D(\text{age})} \quad .$<br>This is the extra risk of dying for individuals with disease that is caused directly by the disease itself. Note that this is different from M<sub>D</sub>, which is the risk of dying from a particular disease for any individual in the population. |
| q<sub>D</sub> | Probability of death due to disease and background mortality. These are not given directly in GBD, but can be calculated using<br>$q_D(\text{age}) = q(\text{age}) - M_D(\text{age}) + em_D(\text{age}) \quad .$<br>Substituting em<sub>D</sub> into q<sub>D</sub> yields<br>$q_D(\text{age}) = q(\text{age}) + M_D(\text{age}) \left( \frac{1}{P_D(\text{age})} - 1 \right) \quad .$ |
|  | We can see that if $P_D=1$, meaning that all individuals in the population have a disease, $q_D$ simply becomes $q$. This is also the case if there is no mortality from disease, so that $M_D=0$. |
| $q_D^{PIM}$ | During the period of increased probability of death due to disease, $q_D$ is used for PIM years. After the period, $q_D$ returns to $q$. |
| $dw$ | Background disability weight from the population overall (0 is no disability and 1 is death).<br>$dw = \frac{YLD_{All\ causes}}{100000}.$ |
| $dw_D$ | Average disability weight due to the disease of interest (0 is no disability and 1 is death).<br>$dw_D(age) = \frac{YLD_D(age)}{P_D(age)}.$ |
| $dw_D^{PID}$ | During the period of increased disability, the disability weight is<br>$dw_D^{PID} = 1 - \left( 1 - \frac{\frac{YLD_{All\ causes}}{100000} - \frac{YLD_D}{100000}}{1 - \frac{YLD_D}{100000}} \right) (1 - dw_D) .$<br>After the period, $dw$ returns to that of the background population ($dw$). |

The GBD database gives the parameters from Table 1 in 5-year age groups to age 95. The under-5 age group is split into “less than one year old” (<1 group) and “1-4 years old”. To obtain single-year age estimates, we undertake the following procedures. For *pop*, we divide the population in the 4- and 5-year age groups evenly by single-year ages, and the terminal age group (95 plus) is divided equally in five parts from 95 to 99. For example, if 500 000 individuals are in the 20-24 age group, we will assume that there are 100,000 20-year olds, 100,000 21-year olds, and so on. For P_D_, I_D_, dw, dw_D_, q(age), and M_D_(age), we assume that the rates (or disability weights) are the same for each single-year age in the aggregate age groups. For example, if dw_D_ was 0.2 in the 20-24 age group, we assume that it was 0.2 for 20-year olds, 0.2 for 21-year olds, and so on.

### Part I: Estimating disease-specific individual life expectancy

The individual life expectancy (iLE) of an individual is simply

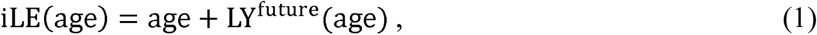

where “age” is the age of the individual and LY^future^ is the number of life years each individual has left to live at their respective age. LY^future^ is not known until the individual actually dies, but its distribution can be estimated as follows. First, we define Y, the maximum age in our lifetable, by setting the chance of surviving from age Y to Y+1 to zero. Then we calculate the number of people who are expected to die at different ages in the Y years to come. This can be done using an upper diagonal (Y+1)× (Y+1) matrix,

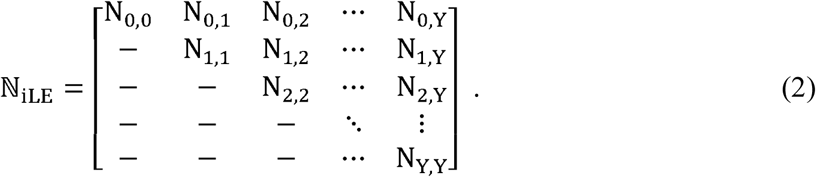

In each element of ℕ_iLE,_ N_c,d_, the *c* denotes the current age and the *d* denotes the age at death. For example, N_3,12_ is the number of today’s 3-year-olds who will die at age 12. Each N_c,d_ is calculated using standard lifetable methodology [22], based on the assumption that q(age) remains the same in the future. In other words, {N_c,O_, N_c,1_,…,N_c,Y_} is the distribution of age at death for an individual with current age *c*. We see that summing the rows, 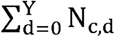, yields the population age structure (pop). Further, summing the columns, 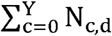, gives the number of people that we expect to die at age *d*. That is, the number of people for which iLE.= *d*. The sum ∑_c_ ∑_d_ N_c,d_ is the total current number of people at all ages. In our calculations, we use Y=99. Note that it follows from the assumption of a static q(age) that average LY^future^ is the same as LE. From (1) we see that iLE is dominated by LY^future^ for young children, and by age for very old people.

Before we break the analysis into diseases, we start by analyzing total figures for one country, as this is familiar for most readers and perhaps more intuitive. Figure 1 shows the distribution of iLE for all age groups in the total population of the United States in 2017. In the left panel, individuals alive in 2017 are ranked by their current age, and the right panel ranks them by iLE. Now we can see that around 22 million people (7%) in the US have iLE < 60 years. As expected, the proportion is much higher in Ethiopia (16%) and Haiti (20%). Note that by simply focusing on LE, we would know nothing about such distributional characteristics.

**Figure 1:**
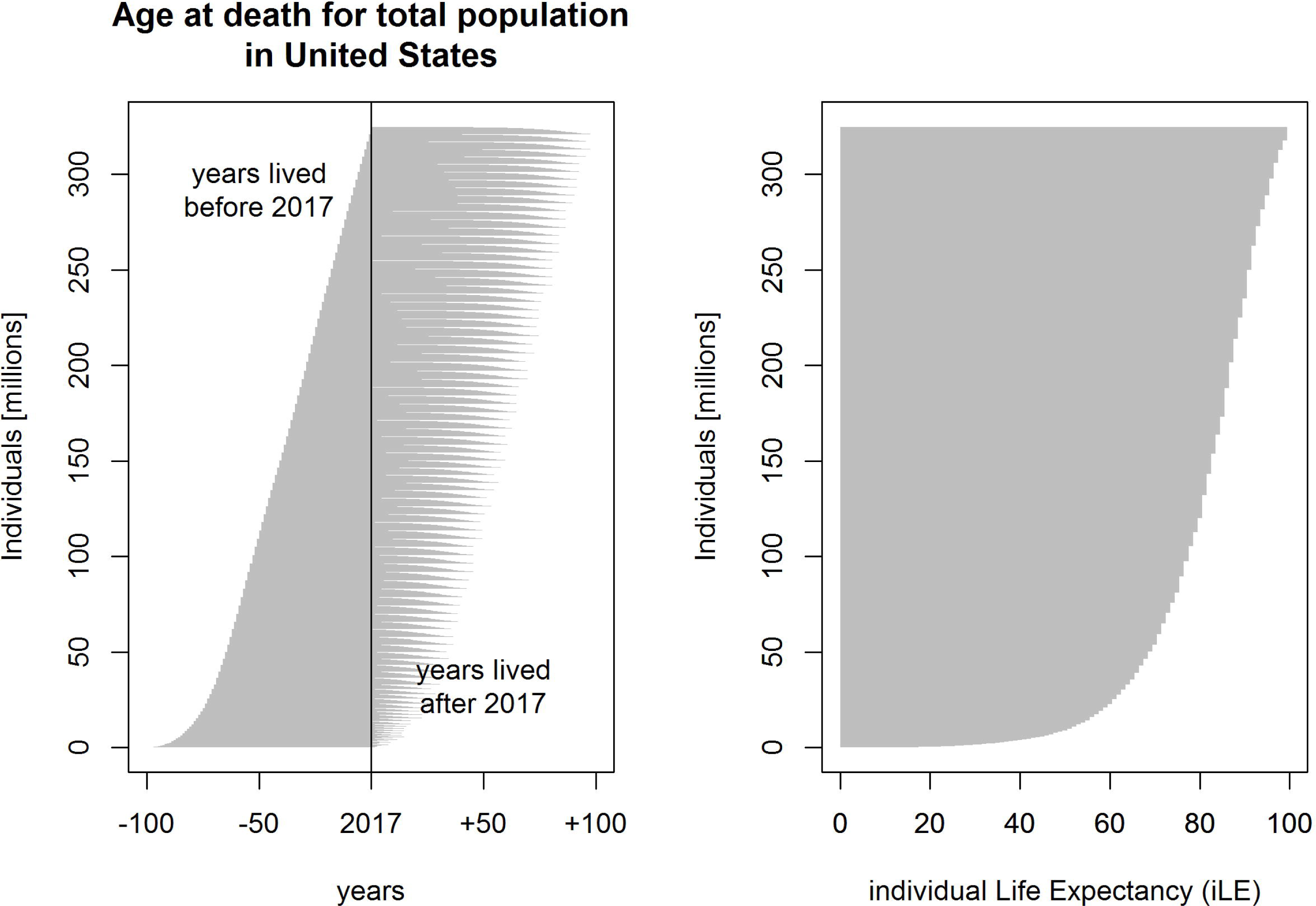
Distribution of individual life expectancy (iLE) for the total US population (2017). Left panel: Distribution by age. On the x-axis, −100 corresponds to the year 1917, and +100 corresponds to the year 2117. Years lived before 2017 are observed, whereas years lived after 2017 are expected. Right panel: Distribution by iLE.

For individuals who get a disease, D, at a particular age, (1) becomes

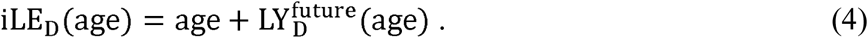

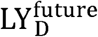 is calculated similarly to LY^future^. Instead of pop(age), we use I_D_(age) × pop (age) (Table 1), and instead of mortality rates for the general population, we use those of individuals with condition D, q_D_ (Table 1). For diseases with very high mortality, 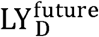 will be small for all ages, and iLE_D_ will therefore to a large extent depend on age alone. If the excess mortality is low, the situation resembles that of (1).

Figure 2 shows the estimated iLE distribution among the 10,600 people with incident cases of acute myeloid leukemia (AML) and 1,950 people with incident cases of acute lymphoid leukemia (ALL) in the United States in 2017. The LE was 67.9 for AML and 67.6 for ALL. However, as the figure shows, the mean age of onset in the United States in 2017 was 62.5 for AML and 45.6 for ALL, and mean 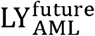 was 5.4 years, whereas 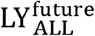 was 22 years. Once again, we see that important information about baseline health is lost when focusing on LE only.

**Figure 2:**
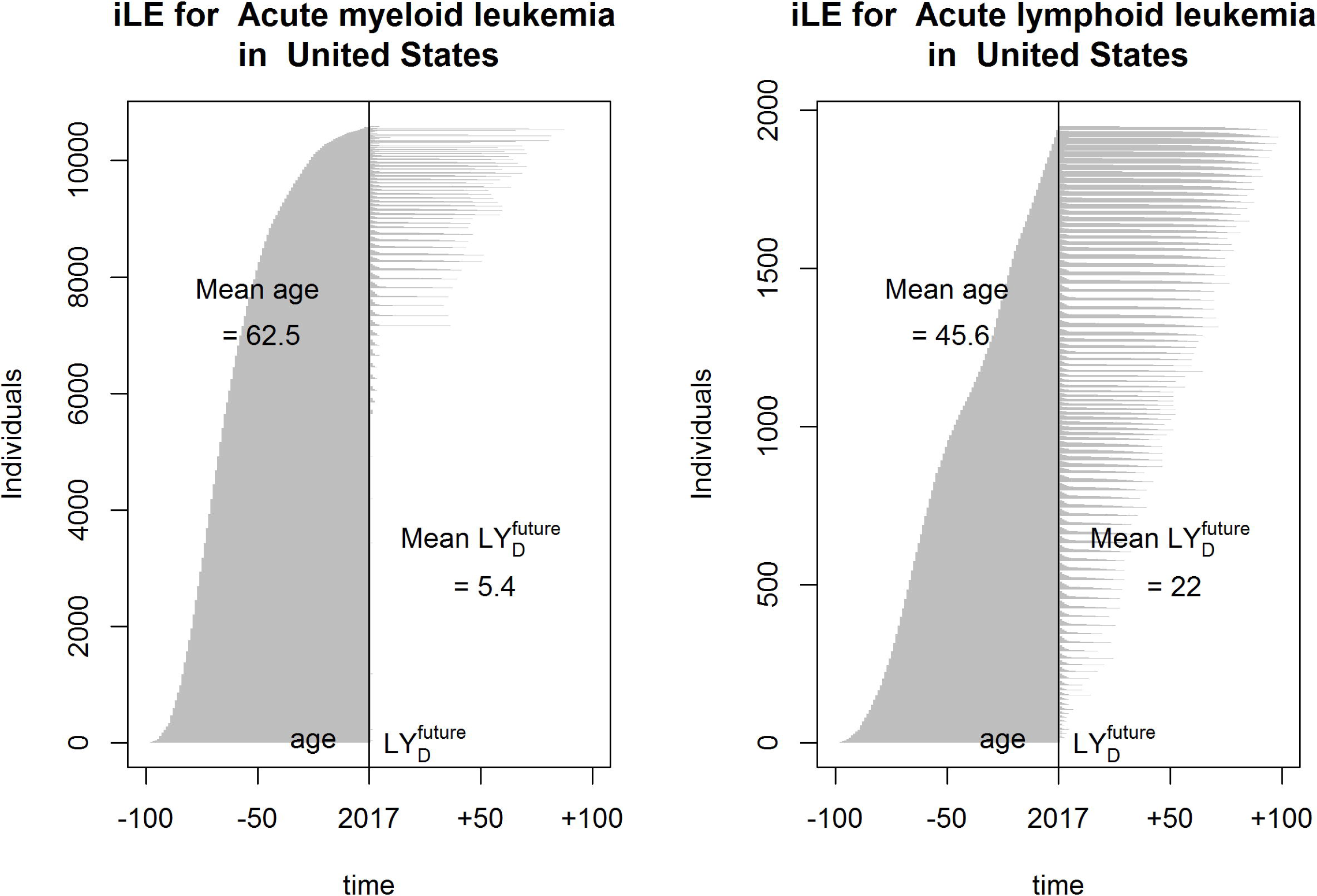
Individual life expectancy (iLE) for individuals who got leukemia in the US in 2017, sorted according to age at onset. Left panel: Distribution for acute myeloid leukemia (AML). Right panel: Distribution for acute lymphoid leukemia (ALL).

### Part II. Adjust for morbidity

Non-fatal morbidities should also be taken into account when assessing baseline health at disease onset, so that one can compare across fatal and non-fatal diseases with different impacts on health loss. This includes estimating health adjusted age (HAA) and future health adjusted life years (HALY^future^). Expanding on (4), we get

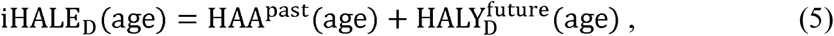

In this section we will explain how to estimate HAA^past^ and 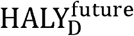 using the background disability, dw, and the excess disease-specific disability, dw_D_ (Table 1).

Figure 3 outlines the conceptual structure of the iHALE method, where both past and future health is summed for each individual with one of the four diseases AML, ALL, epilepsy and schizophrenia.

**Figure 3:**
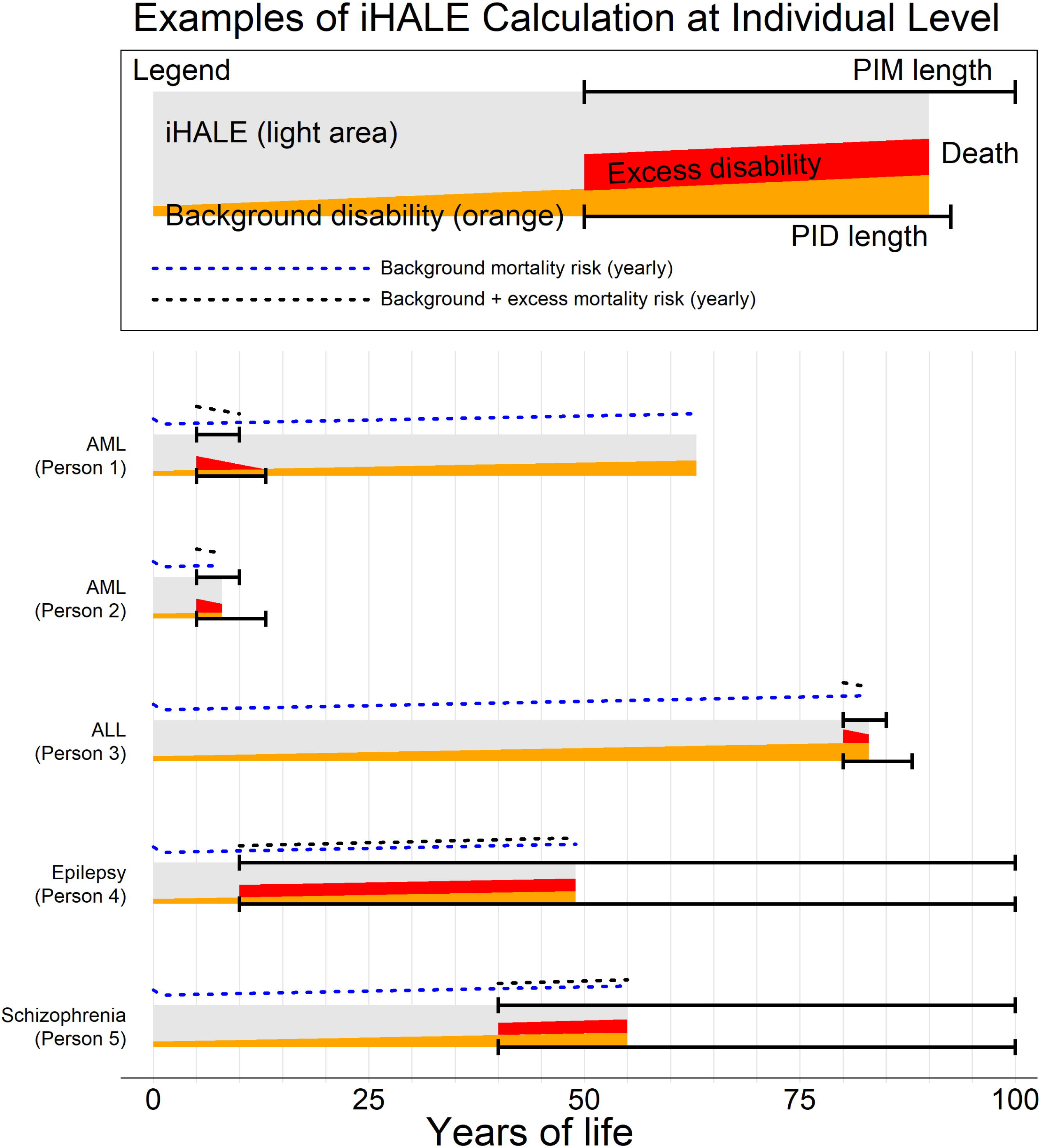
Outline of the conceptual structure of the individual Health Adjusted Life Expectancy (iHALE) method, where we calculate the sum of past health and expected future health for five individuals with different diseases (AML, ALL, epilepsy and schizophrenia). For a disease D, the dashed black line is background mortality, and the blue line is background mortality added to the excess risk of death caused by D. The orange area is background health loss due to disability (dw), and the red area is health loss caused by D (dw_D_). The grey area is iHALE_D_. The sum of the grey, orange and red areas constitute iLE. The top solid black line gives a period after the onset of D when the person had a period of increased mortality (PIM). The bottom line gives a similar period of increased disability (PID).

HAA^past^ is calculated as follows,

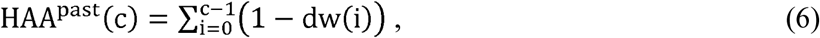

where, *c* is current age and dw(i) is the background disability from age “i” to “i+1” (Table 1). We assume that conditions are independent and that past dw are the same regardless of current disease status.

To account for future non-fatal health loss caused by a disease, D, we use disease specific excess disability, dw_D_, and mortality, q_D_, as calculated in Table 1. However, mortality risk is returned from q_D_ to q after a period of increased mortality (PIM), and morbidity returns from dw_D_ to dw after a period of increased disability (PID) (Table 1 expands on PIM and PID).

In Figure 3 we see from the AML examples how two different persons may fare under the same PIM and PID. Person 1 survives long enough that both mortality and morbidity return to those of the background population, and then dies at age 63 from a different cause, whereas Person 2 dies during the PIM.

Future health adjusted life years, as a function of current age and age at death, is

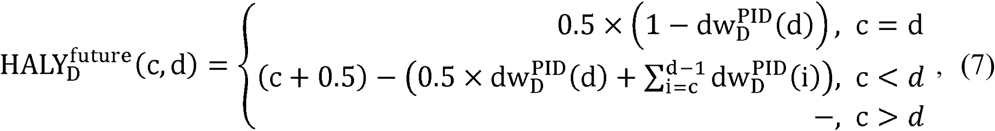

where *c* is current age and *d* is age at death. As in (6), 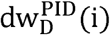 is the disability weight from age “i” to “i+1”. However, because we estimate future health loss, the disability weight must be adjusted during PID. This means that the disability increased for a period of PID years after onset before returning to that of the general population (Table 1).

We next set out to estimate the iHALE distribution in a population. This is done in several steps. First, we create one matrix for past health, and one for future health. The matrix for past health is

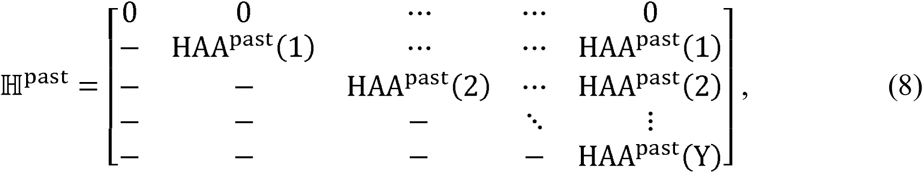

where HAA^past^ is from (6). Because the row number represents current age, we see that the elements are the same within each row. In other words, your past health only depends on your current age, and not your future age at death (column number).

In the matrix for future health, we need to account for both current age and age at death.

Using 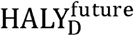 from (7), we get

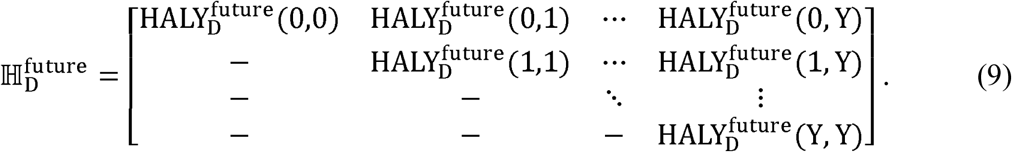

Adding past and future health yields

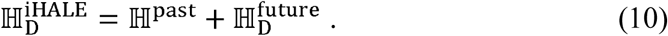

In 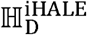, row number *c* gives iHALE for individuals who are *c* years old, whereas column number *d* gives the iHALE for individuals who will die at age *d*. As opposed to the discrete iLE, iHALE is continuous. For example, one individual who dies at age 80 may have an iHALE equal to 67.3 healthy years, whereas another could have achieved 67.4 or 67.5. Because we do not have access to data on individuals, every individual with the same condition and the same age of onset is assumed to have the same iHALE distribution. In ℕ_iLE_ from (2), we obtained these distributions by considering the rows. This information is not available in 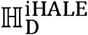, but we may create a new matrix, 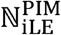, where the elements correspond to those of ℕ_iLE_, but are calculated using 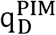 instead of q (Table 1).

Pairing all elements in 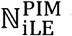 with the corresponding element in 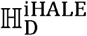 yields the iHALE distribution for all values of *c* and *d* for the disease D.

In 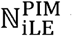 we use incidence to identify those who get the disease D each year, which is especially useful for life-long conditions. See Figure A1 (Appendix) for details on incidence assumptions that are being used in iHALE calculations for AML and ALL in the US.

Figure 4 shows that even though average iHALE is similar for AML (58.7) and ALL (57.7), the distribution across individuals is different. For example, in the US we expect 4.7% of individuals with ALL to have iHALE < 20 (T20), compared with only 2.1% for individuals with AML. Hence, considering risk of a low baseline health, individuals with ALL would be worse off. Still, the 75^th^ percentile (Q3) of iHALE for individuals with ALL (71.3) was higher than in people with AML (67.9), so with respect to chance of a high baseline health individuals with AML would be the worse off. Again, this highlights the need for distributional concerns in policy making.

**Figure 4:**
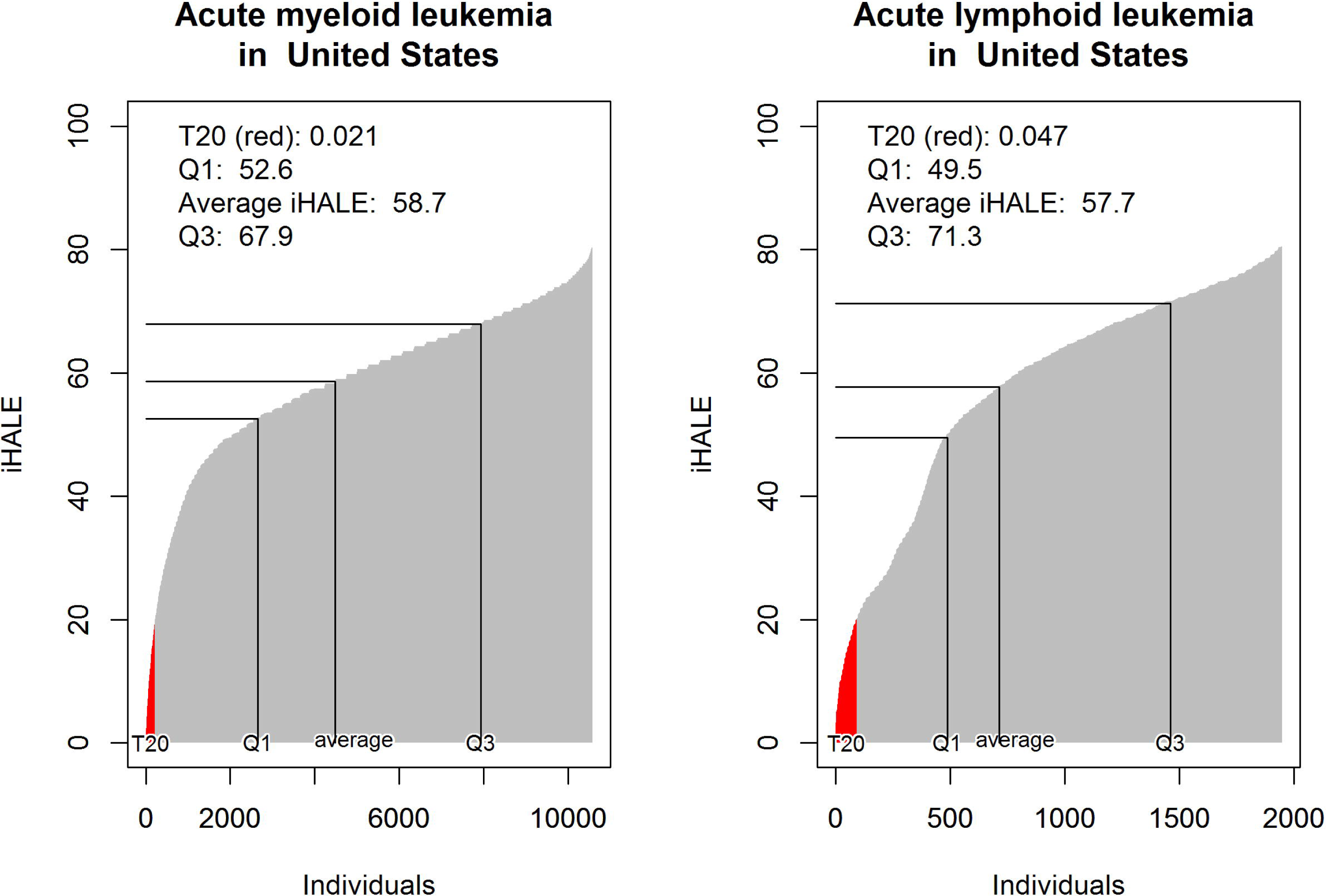
Distribution of individual Healthy Adjusted Life Expectancy (iHALE) for AML and ALL in the United States in 2017.

### Data availability statement

The data used to create the tables and figures in this paper can be accessed without restrictions at https://doi.org/10.5281/zenodo.3258330.

## RESULTS

Table 2 shows average iHALE, T20, Q1 and Q3 for ALL, AML, schizophrenia and epilepsy in six countries (see Appendix Table A4 for 200 NCDI conditions). Rank orders of the four conditions varied both between countries and according to measure of who is worse off.

**Table 2:** Average individual Health Adjusted Life Expectancy (iHALE), T20, Q1 and Q3 for four conditions in six countries, results from 200 NCDI conditions can be found in the Appendix Table A4 (the GBD 2017 study [3] is source for all calculations).

|  |  | Acute lymphoid leukemia | Acute myeloid leukemia | Epilepsy | Schizophrenia |
| --- | --- | --- | --- | --- | --- |
| Ethiopia | Average iHALE | 33.0 | 30.8 | 36.8 | 41.1 |
|  | T20 (%) | 47.5 | 41.2 | 14.1 | 0.5 |
|  | Q1 | 7.8 | 7.0 | 26.3 | 35.6 |
|  | Q3 | 61.0 | 51.7 | 46.2 | 46.0 |
| Haiti | Average iHALE | 33.4 | 33.5 | 39.5 | 40.7 |
|  | T20 (%) | 44.4 | 32.0 | 8.3 | 0.5 |
|  | Q1 | 9.5 | 13.5 | 30.5 | 35.3 |
|  | Q3 | 58.3 | 51.0 | 48.4 | 45.5 |
| China | Average iHALE | 57.5 | 46.5 | 56.6 | 45.7 |
|  | T20 (%) | 5.1 | 15.2 | 1.8 | 0.1 |
|  | Q1 | 47.4 | 31.6 | 50.4 | 39.3 |
|  | Q3 | 71.6 | 62.8 | 65.6 | 51.4 |
| Mexico | Average iHALE | 44.5 | 39.4 | 53.5 | 44.4 |
|  | T20 (%) | 23.7 | 23.0 | 2.0 | 0.2 |
|  | Q1 | 20.4 | 21.4 | 46.8 | 38.7 |
|  | Q3 | 66.8 | 56.6 | 61.7 | 49.7 |
| US | Average iHALE | 57.7 | 58.7 | 58.8 | 43.5 |
|  | T20 (%) | 4.7 | 2.1 | 0.7 | 0.2 |
|  | Q1 | 49.5 | 52.6 | 53.3 | 38.2 |
|  | Q3 | 71.3 | 67.9 | 66.5 | 47.9 |
| Japan | Average iHALE | 66.0 | 62.8 | 64.3 | 48.6 |
|  | T20 (%) | 1.5 | 1.6 | 0.6 | 0.0 |
|  | Q1 | 60.7 | 57.3 | 59.1 | 42.1 |
|  | Q3 | 76.6 | 71.6 | 72.4 | 54.4 |

T20: Proportion of individuals with disease who attain iHALE > 20.
Q1: Attained iHALE for the individual at the 25<sup>th</sup> percentile
Q3: Attained iHALE for the individual at the 75<sup>th</sup> percentile

The iHALE distribution for schizophrenia was similar across the six settings, although Japan stands out in a positive manner. The difference in average iHALE for schizophrenia between the US and Japan was larger than the difference between the US and Ethiopia (Haiti 40.7, Ethiopia 41.1, China 45.7, Mexico 44.4, US 43.5, Japan 48.6). The same applied to the quartiles. T20, the proportion of people with iHALE <20, was low across countries for schizophrenia, which is reasonable, as schizophrenia rarely manifests in childhood. Variability in average iHALE for epilepsy between countries was high (Ethiopia 36.8, Haiti 39.5, Mexico 53.5, China 56.6, US 58.8, Japan 64.3). In addition, there is much more unequal iHALE distribution for epilepsy in countries with low average iHALE (difference between quartiles (Q3-Q1) was 19.9 in Ethiopia, 17.9 in Haiti, 15.2 in China, 14.9 in Mexico, 13.3 in Japan, and 13.2 in the US). Further, only 0.6% of the Japanese had iHALE under 20, but the number was 14.0% among Ethiopians and 8.4% among Haitians.

## DISCUSSION

According to fairness concerns, limited health care resources should be allocated to interventions that benefit the worse off in society [5-7, 18]. In this paper, we present a quantitative method for identifying the worse off by estimating the distribution of lifetime health across individuals in the same disease category. We show how two conditions, ALL and AML, with similar LEs and average iHALEs have substantially different distributions of iHALE. In addition, we show how iHALE varies across countries, and demonstrate how the iHALE distribution captures different aspects of the fact that diseases are typically more severe in low-income than in high-income countries. Our new framework important for priority setting because it can be used to assign extra value to health gains from interventions targeting the worse off. The relevance of iHALE is particularly good for preventive interventions for a disease where you are likely to capture benefits across a range of ages (for example treating strep throat in school children to prevent rheumatic heart disease).

Sullivan, in 1971, suggested how morbidity adjustment could be done for LE to get HALE by modifying the standard life table model to estimate the expected duration of a condition by exposing a birth cohort of a disease specific mortality and disability rate over a lifetime [22]. Sullivan’s method estimates average expected years of healthy life rather than the distribution of iHALE between individuals as done in this paper.

In this article we present iHALE as an achievement measure, but it may be more intuitive to present baseline health at disease onset as a shortfall from what individuals could potentially achieve. iHALE could be converted to an individual gap-measure (e.g. iDALY) by using the YLL method applied in GBD. Shortfall in life years could be calculated for each individual at disease onset by using the lowest mortality by age in the world as a reference. Shortfall in disabilities could use the lowest YLD rates across countries as a reference for disability shortfall. However, disease shortfall measures are beyond the scope of this paper.

PIM and PID, as presented in this paper, have some limitations. They could be different for the same condition across settings, as would be the case for conditions, like HIV, that can be treated or controlled more effectively in some countries than in others. Part of these differences should be captured in the excess mortality differences between countries in our current analysis, but the durations of the periods are also likely to vary. Additionally, PIM and PID do not capture the nature of conditions where mortality and morbidity have complicated temporal patterns. For example, the peak increase in mortality risk for HIV patients is about a decade after onset. At a conceptual level, these obstacles are easy to handle. One simply must estimate PIM and PID for all conditions under consideration in all relevant settings. However, the empirical task of getting precise PIM and PID estimates is not trivial.

Understanding the underlying reasons for differences in the distribution of iHALE can have policy implications. Observed differences in the distribution of iHALE between countries can originate from a number of reasons. It is important to note that as a measure of lifetime health among people with a specific condition, iHALE is influenced by mortality risk and morbidity from other causes, as well as by the age at which the disease occurs. Thus, variation in iHALE could be caused from differences in demography and epidemiology at the country level, or from variations in access to health care that underlie differences in disease-specific morbidity or mortality rates. The relative contribution of each of these differences to the overall difference in iHALE between countries depends on which countries are being compared. How to quantify the role of each factor is discussed further in Appendix (Figure A2).

To measure the true disease-specific baseline health distribution, the past health would be calculated using observed past disability for individuals. The data available for our calculations were limited in several ways. There was no individual-level morbidity and mortality information, so we used population averages. This meant that we were unable to account for correlation between illnesses. For example, people who die of a car accident at age 45 may be different on average from those who die of a myocardial infarction at age 45 regarding lifestyle (smoking, exercise, diet) and biology (metabolism, genetics), which could affect the risk of other morbidities. As a result, our estimates of iHALE may be high (overestimate baseline health) for illnesses that are often experienced with comorbidities because they do not capture the higher burden from the associated illnesses. Conversely, our estimates of iHALE may be low (underestimate baseline health) for illnesses that have few comorbidities. These limitations are especially evident in mental health conditions. The GBD estimates do not attribute any mortality to mental health disorders; however, we know that patients with mental health disorders have higher mortality risk compared to the general population [23-25]. Our schizophrenia iHALE results are additionally limited by distributions of disability weights that do not vary along with treatment availability across countries [26]. The limited time series available from the GBD meant that we did not have complete historical average disability rates. For consistency, we used age-specific rates of disability for the calculations of past health; however, health achievement in a real population would use historical disability information if available.

## CONCLUSION

Increasing availability of demographic and epidemiologic data creates opportunities for quantifying the baseline health at disease onset to guide priority setting in health care. Policy makers, supported by the ethical literature, are concerned about giving higher priority to the worse off. However, the impact of such fairness concerns to health policy does not match the impact cost-effectiveness analysis has had on policy the last decades. Data availability and lack of rigorous methods for quantifying the baseline health at disease onset are likely contributing factors to the negligence of the worse off in de facto health care priority setting. Here we have presented a method for calculating iHALE and illustrate with examples how and why a metric that is sensitive to distribution is relevant for priority setting in health care and the measurement of population health. We lay the foundations for undertaking detailed calculations of disease-specific iHALE in multiple countries.

## LIST OF ABBREVIATIONS

(iHALE): individual Health Adjusted Life Expectancy
(iLE): individual life expectancy
(PIM): Period of Increased Mortality
(PID): Period of Increased Disability
(HAA): health adjusted age
(CEA): cost-effectiveness analysis
(GBD): Global Burden of Diseases
(DALY): disability-adjusted life-years
(HALY): healthy life years
(YLL): years of life lost
(YLD): years lived with disability
(AML): acute myeloid leukemia
(ALL): acute lymphoid leukemia

## Appendix

### A1 Incidence

Figure A1 shows details on incidence assumption that is being used in iHALE calculations.

**Figure A1:**
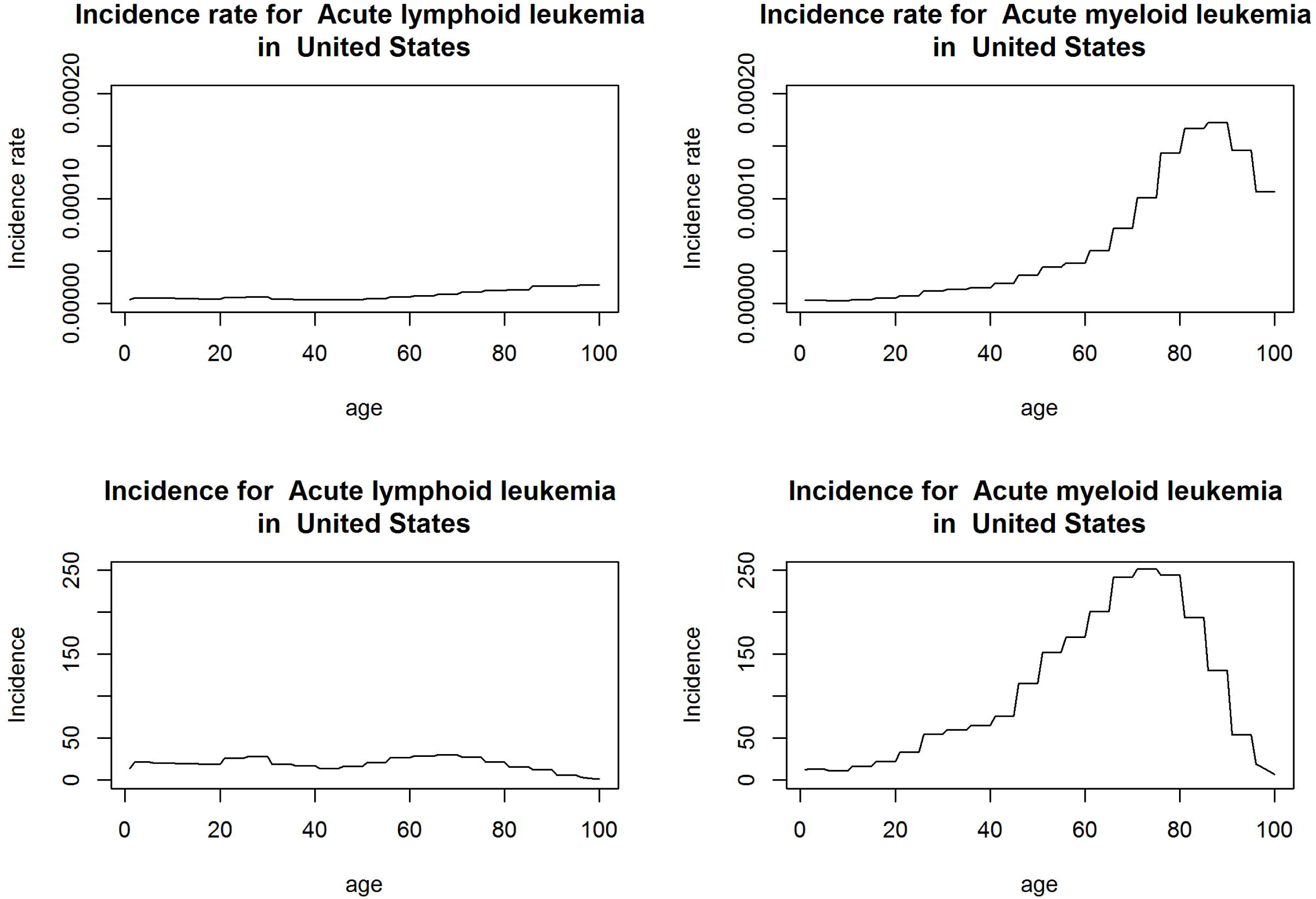
Incidence rates (top) and incidence (bottom) across age groups for AML and ALL in the United States (2017) [1].

### A2 Decomposition

To illustrate how differences in demography, epidemiology, and disease specific mortality and morbidity led to differences in average iHALE between Japan and Ethiopia as exemplars, we took an approach that parallels methods described by Das Gupta [2], creating estimates using the 64 possible scenarios of the two values for the 6 input factors. For each factor, there are then 32 pairs of scenarios in which all five other factors remain constant but the factor in question takes on either Japan or Ethiopia’s value. To calculate the effect of each factor on the overall iHALE difference between Ethiopia and Japan, we took the average of the difference in iHALE between these 32 sets of paired scenarios. Essentially, this is the average effect of varying that one factor with every possible combination of other factors. Figure A2 shows the results of this decomposition. As expected, the younger age structure in Ethiopia and the higher overall mortality contributes substantially to differences in iHALE. However, we also see that disease-specific mortality differences play a large role for epilepsy, leading to a much lower iHALE in Ethiopia. The disease-specific disability plays a very small role for schizophrenia, as discussed in the main text, because of a lack of variation in the distribution of severity across countries in the input data from the GBD data (see A3). Surprisingly, the age distribution of incidence contributes to higher iHALE in Ethiopia, a function of the age distribution of incidence rates from GBD, which tend to be higher at older ages and lower at younger ages in Ethiopia.

**Fig A2:**
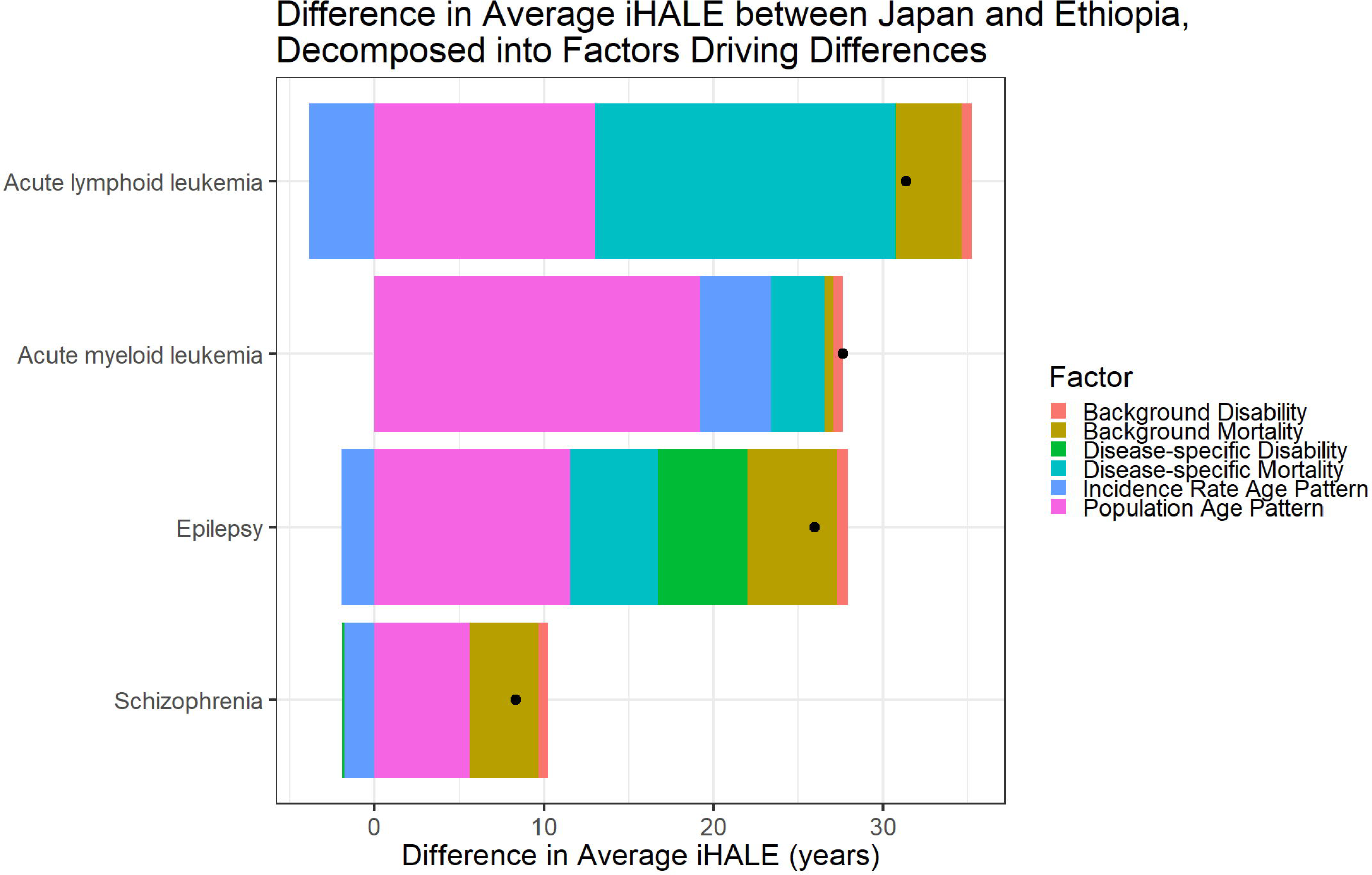
Difference in iHALE between Japan and Ethiopia for 4 diseases (Acute lymphoid leukemia, Acute myeloid leukemia, Epilepsy and schizophrenia), decomposed into factors driving differences.

### A3 Age-standardized average disability weight

Mental disorders like major depressive disorder (MDD), bipolar disorder, and schizophrenia show almost no difference in age-standardized average disability weight between high income and low income countries in GBD, despite higher treatment rates in high income settings. We might expect the distribution of severity to be more severe in low income settings where treatment is less available. Differences in iHALE between high and low income settings, particularly for nonfatal conditions like mental health disorders, are limited by these estimated severities in GBD. Note the much larger difference in average disability weight for epilepsy.

**Figure A3:**
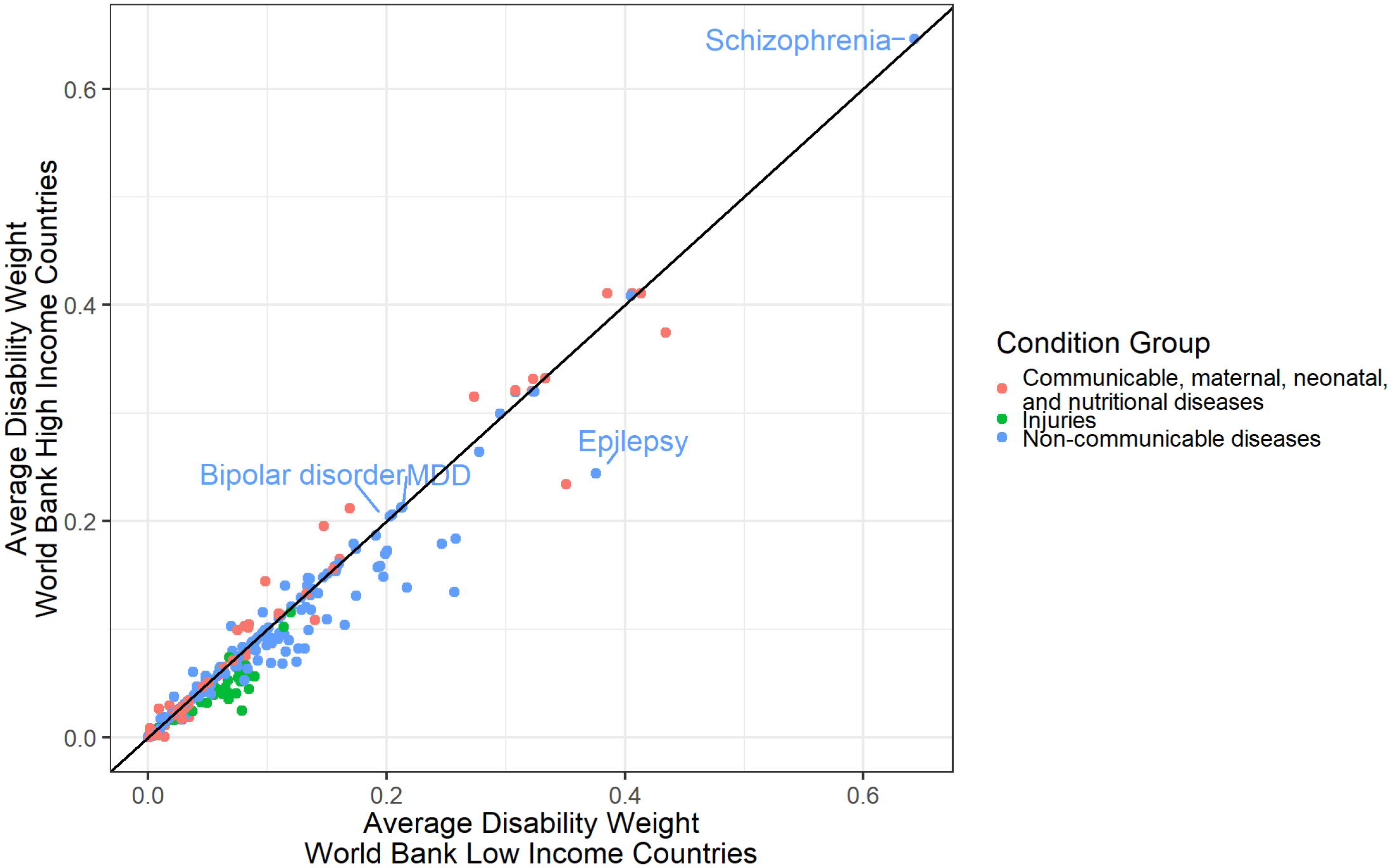
Age-standardized average disability weight (YLDs/Prevalence) in High Income versus Low Income countries by World Bank income groups. MDD=Major depressive disorder

### A4 iHALE for 200 diseases in 6 countries

Table A1 shows results for iHALE for 200 NCDI conditions for Ethiopia, Haiti, China, Mexico, US and Japan.

**Table A1:**
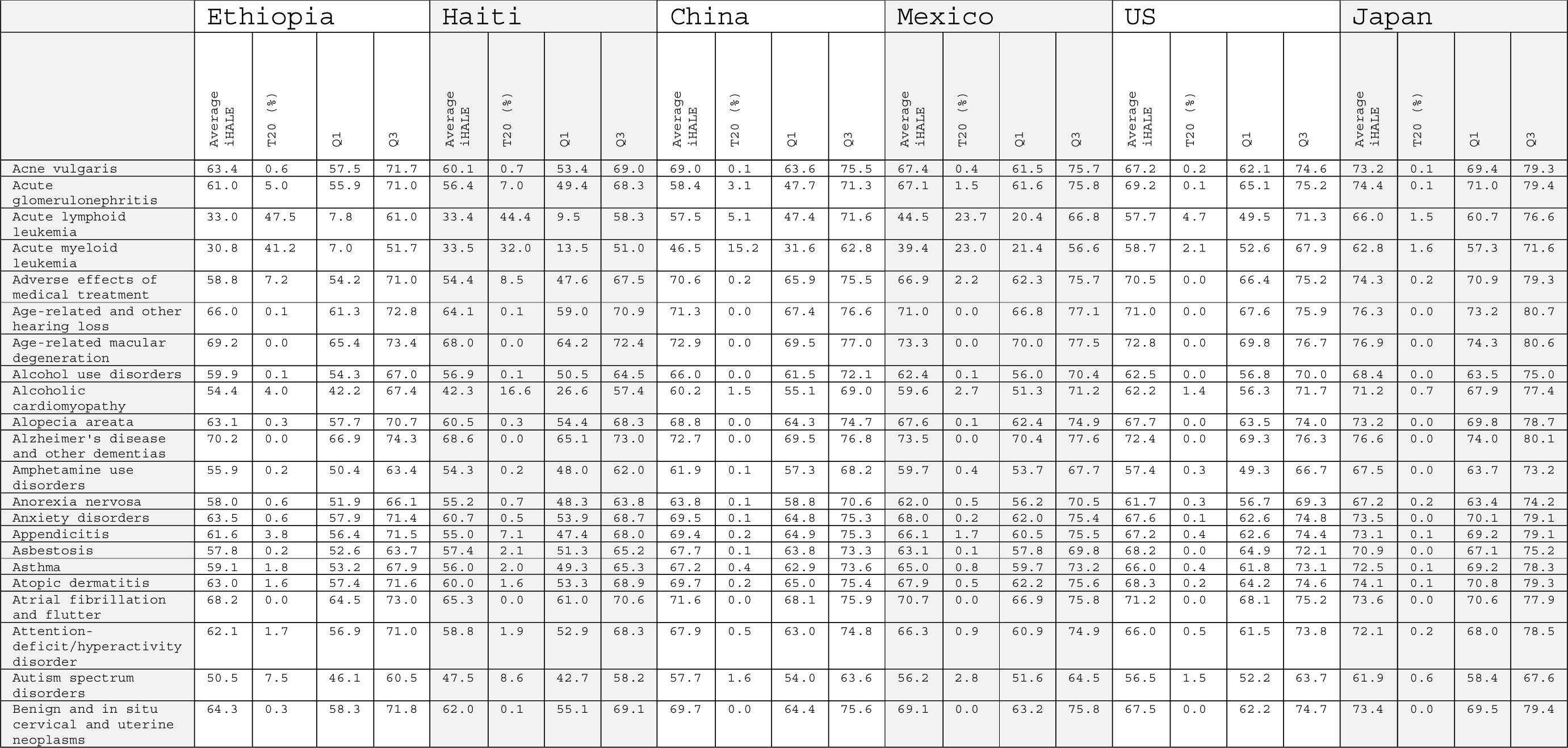
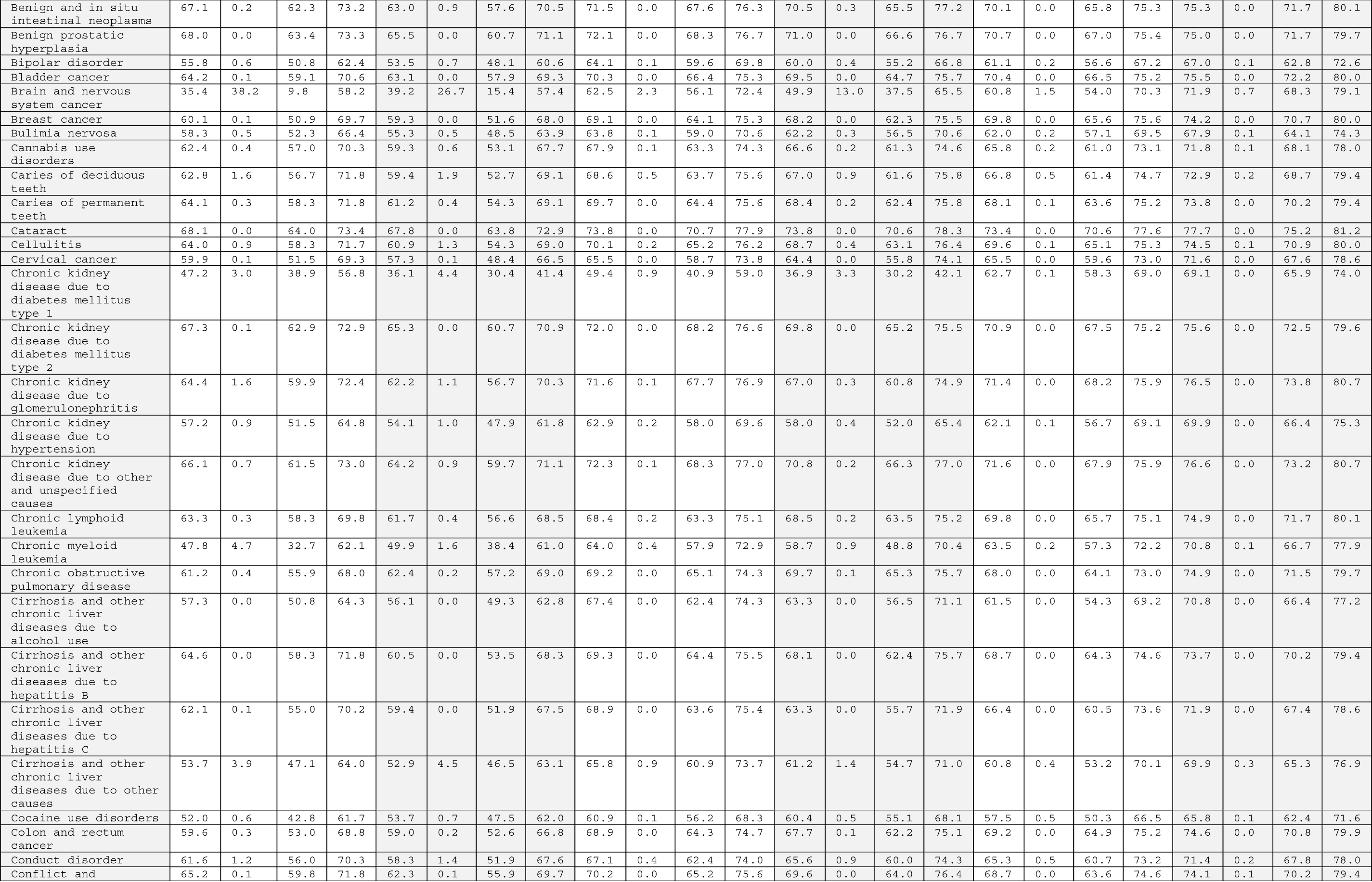
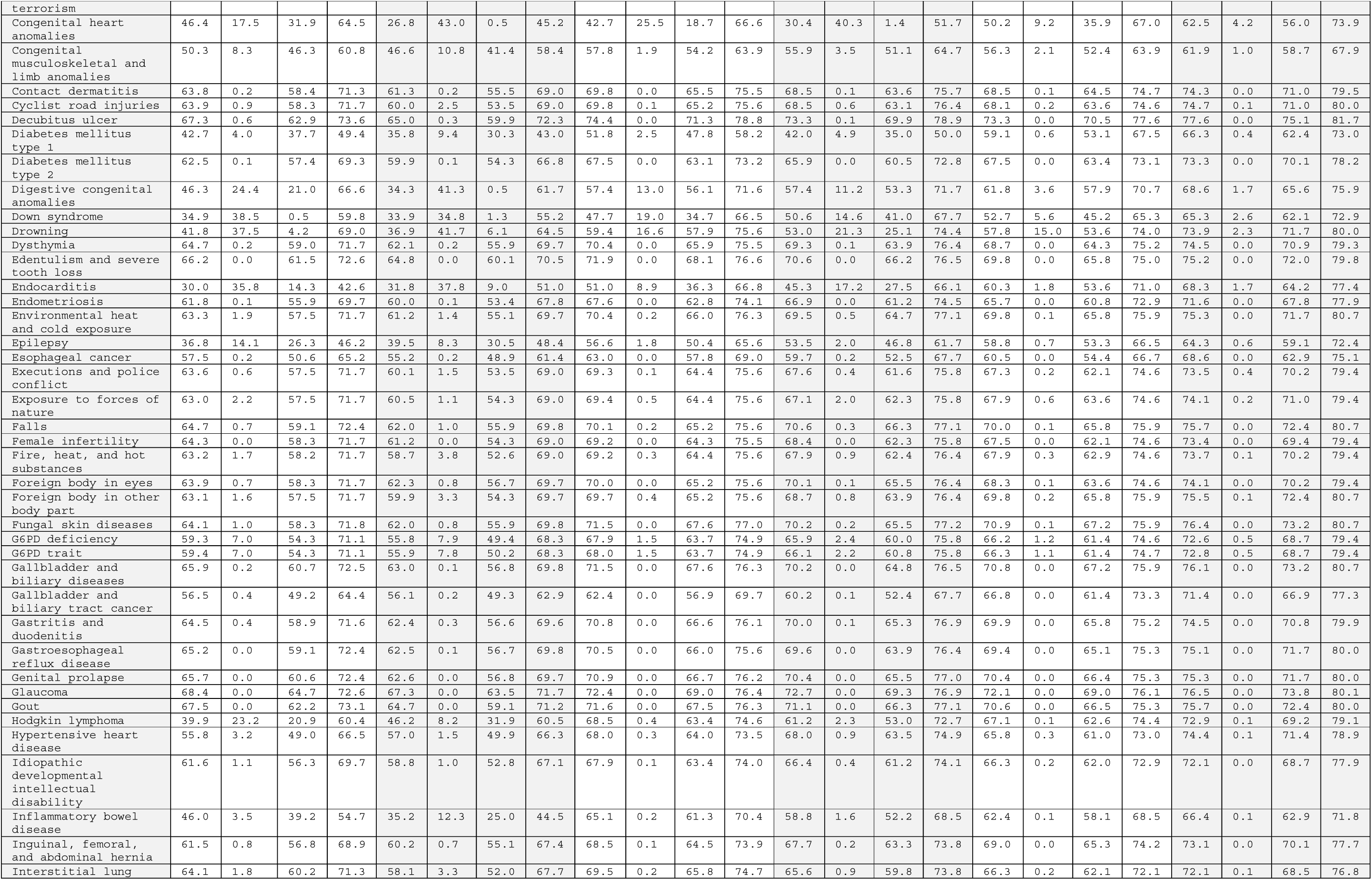
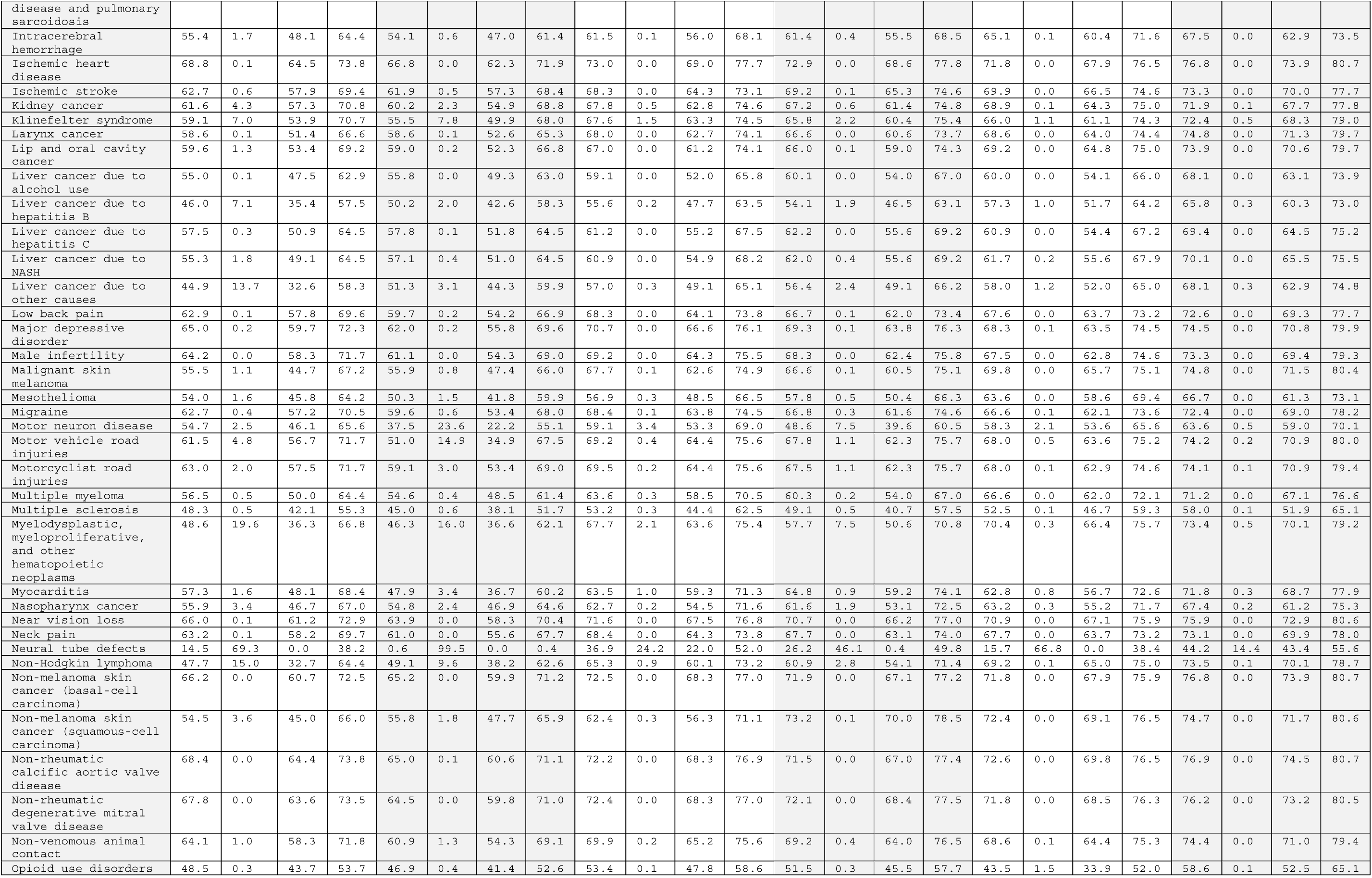
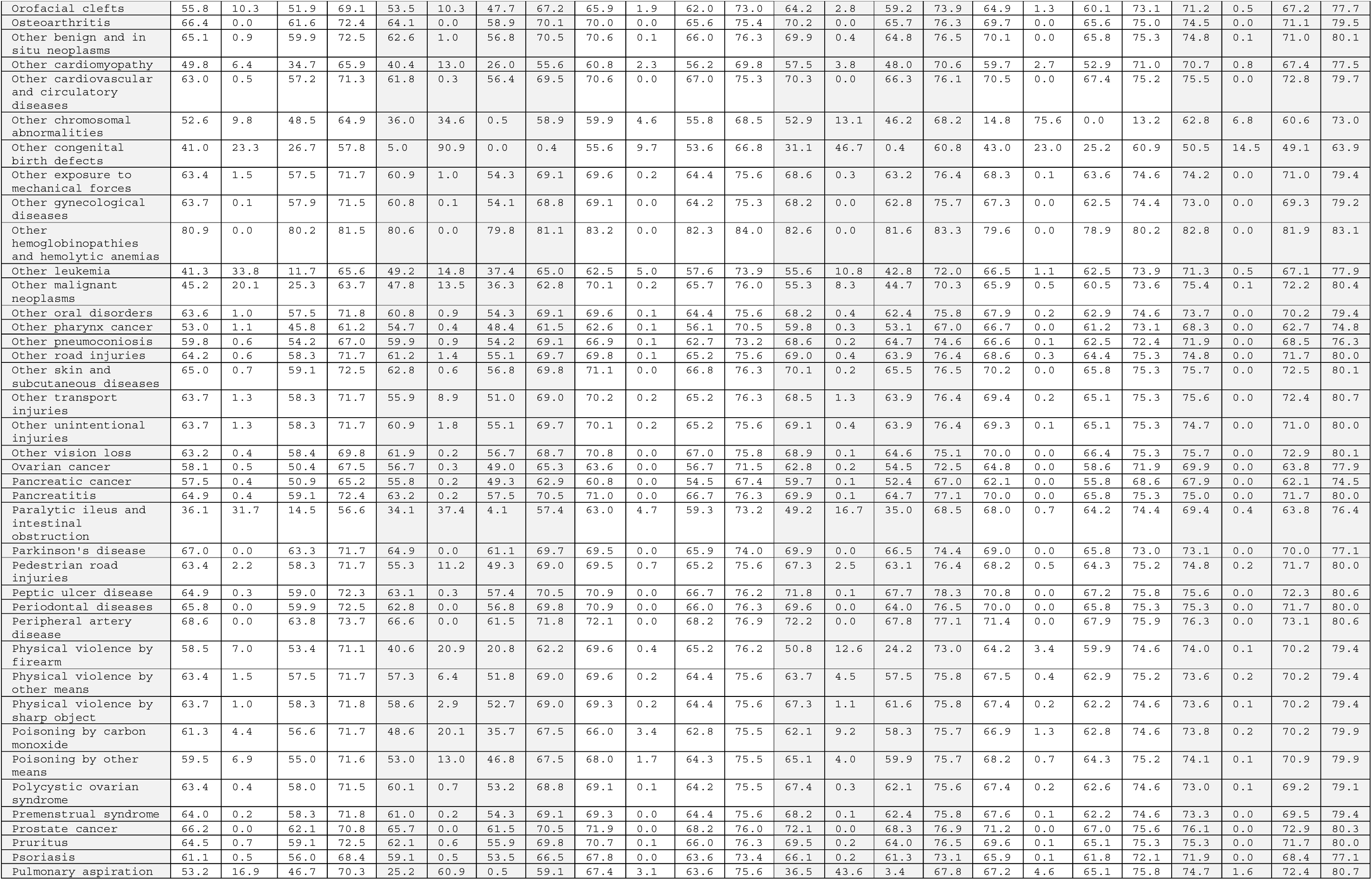
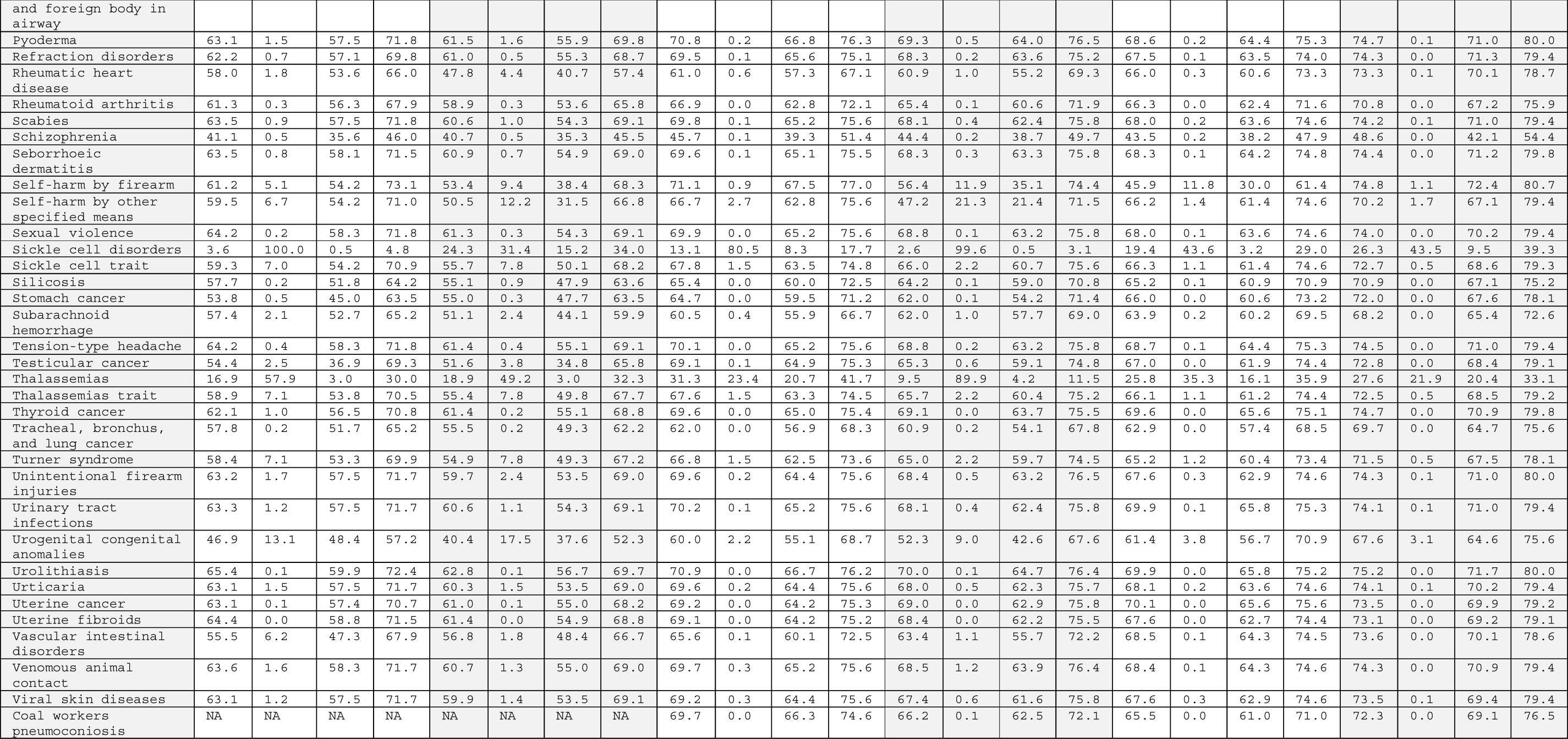
iHALE for 200 NCDI conditions for Ethiopia, Haiti, China, Mexico, US and Japan (T20 is the % with iHALE<20, Q1 is quartile 1 and Q3 is quartile 3).

